# Multi-ancestry Genome-wide Association Study of Inpatient Opioid Exposure Following Knee or Hip Arthroplasty

**DOI:** 10.1101/2025.07.23.25331996

**Authors:** Zeal Jinwala, Christal N. Davis, Jackson F. SooHoo, Million Veteran Program, Joel Gelernter, Rachel L. Kember, Rachel Vickers-Smith, Henry R. Kranzler

## Abstract

Opioids are commonly prescribed to manage acute postoperative pain. However, there is considerable variability in opioid administration patterns and practices, which is partially attributable to patient characteristics. We used electronic health record (EHR) and genotype data from the Million Veteran Program sample to investigate genetic predictors of individual differences in inpatient opioid analgesic exposure following knee or hip arthroplasty (n = 27,896). We extracted data from pharmacy records of administered inpatient opioid medications to derive a measure of average daily opioid exposure during the inpatient postoperative period. We then conducted genome-wide association studies (GWAS) to identify associated genetic variants in individuals of African-like (AFR; n_AFR_ = 4,676), Admixed American-like (AMR; n_AMR_ = 2,126), and European-like (EUR; n_EUR_ = 21,094) genetically inferred ancestries. Models controlled for age, sex, pre-procedure opioid use disorder status, procedure type (knee or hip), length of stay, and the first 10 genetic ancestry principal components. The within-ancestry GWAS were followed by a cross-ancestry GWAS meta-analysis using fixed-effects inverse variance weighting in METAL. No loci reached genome-wide significance in the within- or cross-ancestry GWAS. Five loci were nominally significant (*p* <5 x 10^-6^) in the cross-ancestry GWAS, 9 in the AFR GWAS, 4 in the AMR GWAS, and 3 in the EUR GWAS. This study provides a framework for the use of EHR data to examine the genetics of opioid exposure in postoperative care and indicates the need for larger samples and more precise phenotyping to better understand genetic contributors to individual variation in analgesic requirements.

## Introduction

Opioid analgesics are commonly administered to manage patients’ acute postoperative pain. However, opioid administration patterns vary substantially across individual patients and providers, reflecting both patient-level (e.g., pain sensitivity and prior opioid exposure) and healthcare systems-level factors (e.g., hospital protocols and provider practices) (Burns et al., 2024; Eid et al., 2018; Zanocco et al., 2024). Despite the importance of provider and institutional practices, a recent estimate suggests that up to 79% of the variability in intraoperative opioid dosing may be attributable to patient characteristics, indicating that opioid administration patterns capture meaningful interindividual variation. Despite this, little is known about the genetic predictors of individual differences in opioid administration in acute postoperative care settings.

A better understanding of the risk factors that contribute to greater postoperative opioid exposure could help optimize perioperative pain management while minimizing opioid use disorder (OUD) risk. In the United States between 1990 and 2010 the drive to treat pain aggressively produced a sharp increase in prescription opioid exposure, one factor contributing to the epidemic of opioid misuse, dependence, and overdose deaths (Okie, 2010). In the Veterans Health Administration, opioid prescribing nearly doubled from 2004 to 2012 (Mosher et al., 2015) and was accompanied by a substantial rise in opioid-involved overdose deaths (Bohnert et al., 2014). Despite recent reductions in opioid prescribing and overdose deaths, there is a need for empirically based, individualized opioid prescribing, particularly in acute care settings where individuals are often first exposed to opioids (Ladha et al., 2019).

Genetic studies of opioid response and postoperative opioid exposure have been limited. Candidate gene studies have yielded inconsistent findings (Coulbault et al., 2006; Fukuda et al., 2009; Nishizawa et al., 2009), and genome-wide association studies (GWAS) have typically been conducted in small samples (*n*s<650) (Cook-Sather et al., 2014; Nishizawa et al., 2014). These GWAS have implicated genes such as *CREB1*, *TAOK3*, and *TRPC3*, but these findings remain to be replicated. A recent GWAS of the codeine prescription count in a sample of 8,639 patients identified 9 genome-wide significant (GWS) loci and a significant association with genetic liability for OUD (Song et al., 2024). However, that study focused on a single opioid and included only European-like (EUR) genetic ancestry individuals. Across studies, genetic variants implicated in post-surgical pain and their potential influence on opioid sensitivity have shown largely weak and inconsistent associations, with limited reproducibility. Thus, larger GWAS of opioid sensitivity in multiple genetically inferred ancestry (GIA) groups are needed to identify genetic predictors of opioid dosing for postoperative pain management (Frangakis et al., 2023).

Electronic health record (EHR)-linked biobanks provide an opportunity to examine genetic predictors of real-world opioid exposure patterns in large and diverse samples. Using EHR data from the Million Veteran Program (MVP), we investigated individual differences in opioid analgesic dosing following knee or hip arthroplasty. These common orthopedic procedures typically cause moderate-to-severe postoperative pain (Chan et al., 2013; Højer Karlsen et al., 2015), making them well-suited for studying the genetic variation that underlies opioid dosing. A study examining perioperative prescribing patterns following total knee arthroplasty showed that patients taking short-acting opioids after surgery had 38% higher readmission risk than those not taking opioids—likely due to accompanying complications, suggesting a need for improved perioperative opioid prescribing strategies (Mudumbai et al., 2020). We focused on opioid medications administered during an inpatient stay and derived a measure of average daily opioid exposure. Our aim was to characterize the genetic predictors of inpatient opioid exposure intensity following arthroplasty. Examining opioid exposure patterns in a large, diverse healthcare system provides a framework for understanding biological contributions to variability in acute postoperative pain management in real-world clinical settings.

## Methods

### Sample

Data were obtained from participants in the MVP, a large biobank comprising U.S. veterans who gave informed consent and provided a blood sample for DNA extraction and genotyping and access to their EHR. Study procedures were approved by the VA Central Institutional Review Board (IRB) and each local IRB. We selected patients who underwent knee or hip arthroplasty procedures between 2000 and 2023 using International Classification of Diseases (ICD)-9 and ICD-10 diagnoses and Current Procedural Terminology (CPT) codes (Supplementary Table 1). For patients who underwent multiple arthroplasties during the study period, we included only the first procedure in the analysis to ensure independent observations.

### Phenotyping

*Opioid Exposure.* Our GWAS phenotype was the average daily opioid exposure during hospitalization. Data on inpatient opioid administration during the arthroplasty hospitalization were extracted from VA pharmacy records using drug class codes “CN101” and “RE301” from the National Drug Code classification system. However, dose unit and formulation information needed to estimate morphine milligram equivalents (MME) were not consistently available, as evidenced by 75.79% of hospitalizations, which included at least one opioid administration whose recorded units or formulation could not be accurately converted using MME (Supplementary Table 2). Because dropping these opioid administrations or excluding the stays entirely would have either biased opioid exposure downward or substantially reduced the analytic sample, we constructed a proxy measure. This measure was based on the number of doses for each opioid administration, scaled by the drug-specific potency factors from an MME conversion table (https://pain.ucsf.edu/opioid-analgesics/calculation-oral-morphine-equivalents-ome), and averaged across the inpatient stay. Specifically, we used the following equation to estimate opioid exposure intensity:

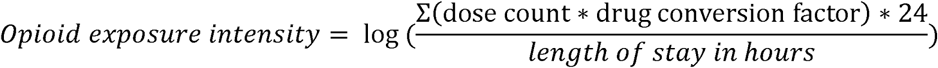

The resulting phenotype was positively skewed (M = 68.39, SD = 148.96) and was thus log-transformed.

We compared this opioid exposure phenotype to a conventional MME measure obtained in the subset of hospitalizations (n = 6,755) where all necessary details on opioid administrations were available. The two measures were moderately to strongly correlated (Pearson r = 0.64; Spearman ρ = 0.75; *p*s < 0.0001), supporting use of the proxy measure to capture inter-individual variation in inpatient opioid exposure.

*Opioid Use Disorder Status.* Pre-existing OUD was defined using ICD-9 and ICD-10 diagnosis codes documented in the EHR prior to the arthroplasty procedure (Supplementary Table 2). Individuals were classified as having OUD if they had at least one inpatient or outpatient diagnostic code consistent with opioid abuse, dependence, or use disorder before the surgery date. OUD status was modeled as a binary covariate in all GWAS to account for potential differences in opioid tolerance and prescribing patterns.

### Genotyping and Imputation

Samples were genotyped using a custom Affymetrix Axiom Biobank Array. Quality control and imputation were performed by the MVP working group (Hunter-Zinck et al., 2020). Participants whose genetic and phenotypic sex did not match, who had seven or more relatives in MVP (kinship > 0.08), or who demonstrated excessive heterozygosity or a genotype call rate <98.5% were removed from the analysis. Monomorphic variants, variants with a Hardy-Weinberg equilibrium P-value <1 x 10^-6^ or a call rate <0.95 were removed. Genotypes were phased using SHAPEIT4 (Delaneau et al., 2019) and imputed using Minimac4 software (Fuchsberger et al., 2015). A combination of the 1000 Genomes Phase 3 panel and the African Genome Resources reference panel was used to impute biallelic SNPs. GIA assignments were determined using the smartpca module in the EIGENSOFT package (Patterson et al., 2006; Price et al., 2006), as outlined in previously (Verma et al., 2022).

### Genome-wide association studies

GWAS were conducted separately within European-like (EUR; n_EUR_ = 21,094), African-like (AFR; n_AFR_ = 4,676), and Admixed American-like (AMR; n_AMR_ = 2,126) individuals. Linear regression models were implemented using PLINK v.2.0, with procedure type (knee or hip), age, sex, pre-existing OUD diagnosis, length of stay (in hours), and the first 10 within-ancestry genetic principal components (PCs) included as covariates. Although our opioid exposure phenotype was normalized per day, we included length of stay as a covariate in the GWAS models to reduce confounding by procedure-related factors (e.g., surgical complexity or recovery trajectory) that could influence opioid prescribing.

Variants with an imputation quality (INFO) score <0.8 were excluded. A minor allele count (MAC) threshold of 100 was applied to the AFR and AMR GWAS because of their smaller sample sizes, with a MAF of 1% used for the EUR GWAS. To account for relatedness, we randomly removed one individual from each pair of related individuals. The within-ancestry GWAS summary statistics were meta-analyzed using a fixed-effects inverse-variance weighted approach in METAL (Willer et al., 2010). To help avoid spurious associations, we required that SNPs were present in at least 5,000 individuals to be included in the meta-analysis. This yielded in 4,899,645 SNPs in the cross-ancestry meta-analysis.

Lead SNPs were identified using a two-step linkage disequilibrium (LD) clumping process on the Functional Mapping and Annotation of Genome-Wide Association Studies (FUMA) website (Watanabe et al., 2017). First, SNPs with P-value ≤5 x 10^-6^ and independent at r^2^ <0.6 were selected as independent GWS SNPs, with lead SNPs defined as those independent from each other at r^2^ <0.1, and candidate SNPs defined as those in LD (r^2^ ≥0.6) with lead SNPs and with a P-value ≤0.05. We used the 1000 Genomes reference panel (Auton et al., 2015) to estimate the r^2^ between SNPs. Nominally significant lead SNPs were identified using a similar two-step process, but with 5 x 10^-6^ as the P-value threshold. Lead SNPs were mapped to genes using positional proximity. We estimated SNP-based heritability of the GWAS using a C++ implementation of linkage disequilibrium score regression (LDSC) (Bulik-Sullivan et al., 2015) available at https://github.com/ykhan1999/ldsc-cpp.

## Results

The sample comprised 27,896 individuals, was predominantly male (92.65%), and had a mean (SD) age of 66.04 (9.01) years. Overall, 4.15% of participants had a pre-existing diagnosis of OUD. The mean of the log-transformed opioid exposure intensity variable was 3.12 (SD = 1.43). See Table 1 for sample characteristics broken down by procedure type and GIA group.

**Table 1.** Sample Characteristics.

|  | **Arthroplasty**<br>**(n = 27,896)** | **Knee Arthroplasty**<br>**(n = 18,534)** | **Hip Arthroplasty**<br>**(n = 9,362)** |
| --- | --- | --- | --- |
|  | Mean (SD) or % (N) | Mean (SD) or % (N) | Mean (SD) or % (N) |
| Age (years) | 66.04 (9.01) | 66.00 (8.63) | 66.12 (9.74) |
| Male | 92.65% (25,847) | 92.20% (17,088) | 93.56% (8,759) |
| Opioid use disorder | 4.15% (1,159) | 3.78% (700) | 4.90% (459) |
| Log (MME_proxy) | 3.12 (1.43) | 3.18 (1.37) | 3.00 (1.52) |
| **African-like** | **N = 4,676** | **N = 2,847** | **N = 1,829** |
| Age (years) | 62.18 (8.63) | 62.31 (8.29) | 61.97 (9.13) |
| Male | 90.21% (4,218) | 88.51% (2,520) | 92.84% (1,698) |
| Opioid use disorder | 7.85% (367) | 7.45% (212) | 8.47% (155) |
| Log(MME proxy) | 3.10 (1.40) | 3.18 (1.35) | 2.98 (1.45) |
| **Admixed American-like** | **N = 2,126** | **N = 1,604** | **N = 522** |
| Age | 64.22 (9.39) | 64.40 (9.07) | 63.67 (10.30) |
| Male | 94.21% (2,003) | 94.01% (1,508) | 94.83% (495) |
| Opioid use disorder | 5.50% (117) | 4.93% (79) | 7.28% (38) |
| Log (MME proxy) | 3.11 (1.39) | 3.15 (1.36) | 2.97 (1.48) |
| **European-like** | **N = 21,094** | **N = 14,083** | **N = 7,011** |
| Age | 67.08 (8.79) | 66.93 (8.41) | 67.38 (9.51) |
| Male | 93.04% (19,626) | 92.74% (13,060) | 93.65% (6,566) |
| Opioid use disorder | 3.20% (675) | 2.90% (409) | 3.79% (266) |
| Log (MME proxy) | 3.12 (1.44) | 3.18 (1.38) | 3.01 (1.54) |

In the cross-ancestry GWAS meta-analysis, no SNPs were GWS. Five independent loci reached the nominal significance threshold (*p* < 5e-6) (Table 2), mapping to three protein-coding genes (Supplementary Table 3). The SFXN3 (Sideroflexin 3) gene encoding a mitochondrial membrane transporter crucial for cellular metabolism and synaptic health was among these functionally mapped genes. Three of the SNPs were found only in EUR individuals, while two (rs72704678 and rs6536582) were present in the GWAS of all three ancestry groups, showed the same direction of effect across groups, and were not significantly heterogeneous in effect size (*p*s > 0.11).

**Table 2.** Summary of lead single-nucleotide polymorphisms reaching nominal significance (p < 5 x 10-6) in the cross-ancestry genome-wide association study meta-analysis.

| **SNP** | **CHR** | **Position** | **A1** | **A2** | **P-value** | **MAF** | **Nearest Gene** |
| --- | --- | --- | --- | --- | --- | --- | --- |
| rs113672700 | 10 | 102807611 | T | C | 5.39E-07 | 0.03 | *SFXN3* |
| rs72704678 | 14 | 95473868 | T | G | 2.89E-06 | 0.91 | *RP11-991C1.2* |
| rs141284882 | 4 | 58777959 | CA | C | 2.98E-06 | 0.89 | *RP11-4O3.2* |
| rs6536582 | 4 | 162138234 | A | G | 3.85E-06 | 0.25 | *RP11-234O6.2* |
| rs117257541 | 13 | 113012499 | T | C | 4.62E-06 | 0.02 | *SPACA7* |

### Within-ancestry GWAS

In the AFR GWAS, no loci were GWS. Nine independent loci reached the nominal significance threshold (Supplementary Table 4). Functional annotation mapped 62 protein-coding genes to the nominally significant loci (Supplementary Table 5). *LYPD6*, whose protein produce modulates nicotinic acetylcholine receptor signaling, was among the functionally mapped genes, as was *ALDH1B1*, a member of the aldehyde dehydrogenase family. No clear enrichment for genes previously implicated in opioid pharmacology or nociception was observed. In the AMR GWAS, no loci reached genome-wide significance, though four loci met the nominal threshold (Supplementary Table 6), mapping to 13 protein-coding genes (Supplementary Table 7). SNP-based heritability was 0.0621 (0.1837) and −0.1474 (0.2499) for AFR and AMR, respectively. In the EUR GWAS, no loci were GWS, though three loci were nominally significant (Supplementary Table 8). These mapped to two protein-coding genes that were also implicated by the cross-ancestry GWAS (*SFXN3* and *SPACA7*) (Supplementary Table 9). SNP-based heritability of the EUR GWAS was 0.0464 (0.027).

## Discussion

This study is the first genome-wide investigation of variation in inpatient opioid exposure following knee or hip arthroplasty. Leveraging data from the MVP, a large and ancestrally diverse biobank with linked EHR data, we identified 27,896 individuals who underwent one of these surgeries for inclusion in GWAS. Although no GWS loci were identified, the approach used advances the field beyond traditional candidate gene approaches (Candiotti et al., 2011; Coulbault et al., 2006; Fukuda et al., 2009; Kumar et al., 2011; Nishizawa et al., 2009) and small-sample GWAS (Aoki et al., 2015; Cook-Sather et al., 2014; Nishizawa et al., 2014) that previously used to study these effects.

Postoperative opioid exposure is likely influenced by biological, clinical, and administrative factors. Although genetic variation contributes to differences in pain sensitivity, opioid tolerance, and pharmacologic responses, inpatient opioid exposure also reflects factors like provider decision-making, institutional protocols, surgical complexity, and prescribing guidelines. Thus, these factors may attenuate genetic effects or make them more difficult to detect. The absence of genome-wide significant findings here may reflect the highly polygenic architecture or substantial non-genetic influences on inpatient opioid exposure.

Several study limitations warrant consideration. First, opioid exposure was estimated using a proxy measure of dose counts and drug-specific potency factors rather than complete MME data. Although the proxy was moderately-to-strongly correlated with MME estimates in a validation subset of hospitalizations, it likely included measurement error that impacted our ability to detect genetic associations. Second, changes in opioid prescribing patterns and provider preferences may have introduced heterogeneity in opioid exposure that was not fully captured in our GWAS models. Thus, studies that consider these factors’ effects on opioid dosing are needed. Collider bias may also have influenced our findings, insofar as all individuals in our sample underwent arthroplasty. Thus, the genetic associations that were identified as nominally significant may reflect factors related to the need for an arthroplasty in addition to opioid dosing requirements. Finally, the MVP is predominantly comprised of male veterans, limiting the generalizability of the findings to other populations.

Despite these limitations, this study demonstrates the feasibility of using EHR-derived inpatient pharmacy data to examine the genetic architecture of real-world opioid exposure. Leveraging a large, multi-ancestry biobank provides a foundation for future genomic investigations of postoperative pain management. Continued efforts that incorporate larger samples and improved phenotyping of opioid dosing will be critical for advancing precision pain management approaches.

## Data Availability

The cross-ancestry and within-ancestry GWAS and meta-analysis summary-level association data will be available in the database of Genotypes and Phenotypes (dbGaP) (https://www.ncbi.nlm.nih.gov/gap/) under accession phs001672 "Veterans Administration (VA) MVP Summary Results from Omics Studies." Registration and approval are needed following dbGaP's data access process.

## Acknowledgements

The views expressed in this article are those of the authors and do not necessarily represent the position or policy of the Department of Veterans Affairs or the US Government. The MVP Core Acknowledgment is in Supplementary Material. This research is based on data from the Million Veteran Program, Office of Research and Development, Veterans Health Administration. The work was supported by Merit Review Awards I01 CX001734 and I01 BX004820 (to HRK); Career Development Award IK2 RD001004 (to CND); the VISN 4 Mental Illness Research, Education and Clinical Center (to CND, RLK, and HRK); and National Institutes of Health grant K01 AA028292 (to RLK). The authors thank the Million Veteran Program staff, researchers, and volunteers who have contributed to MVP, and especially those who served their country in the military and agreed to enroll in the study (mvp.va.gov). Scott Damrauer, M.D. provided helpful comments on the manuscript.

## Author Contributions

ZJ, CND, JFS, and RVS conducted data analyses. ZJ and CND drafted the manuscript. JG, RVS, RLK and HRK reviewed and provided substantive comments on the manuscript. HRK conceived the project, obtained funding to support it, and supervised the project. All authors reviewed and approved the final version of the manuscript.

## Data availability

The within-ancestry GWAS and cross-ancestry meta-analysis summary-level association data are available in the database of Genotypes and Phenotypes (dbGaP) (https://www.ncbi.nlm.nih.gov/gap/) under accession phs001672 “Veterans Administration (VA) MVP Summary Results from Omics Studies.” Registration and approval are needed through dbGaP’s data access process.

---

**Figure 1:**
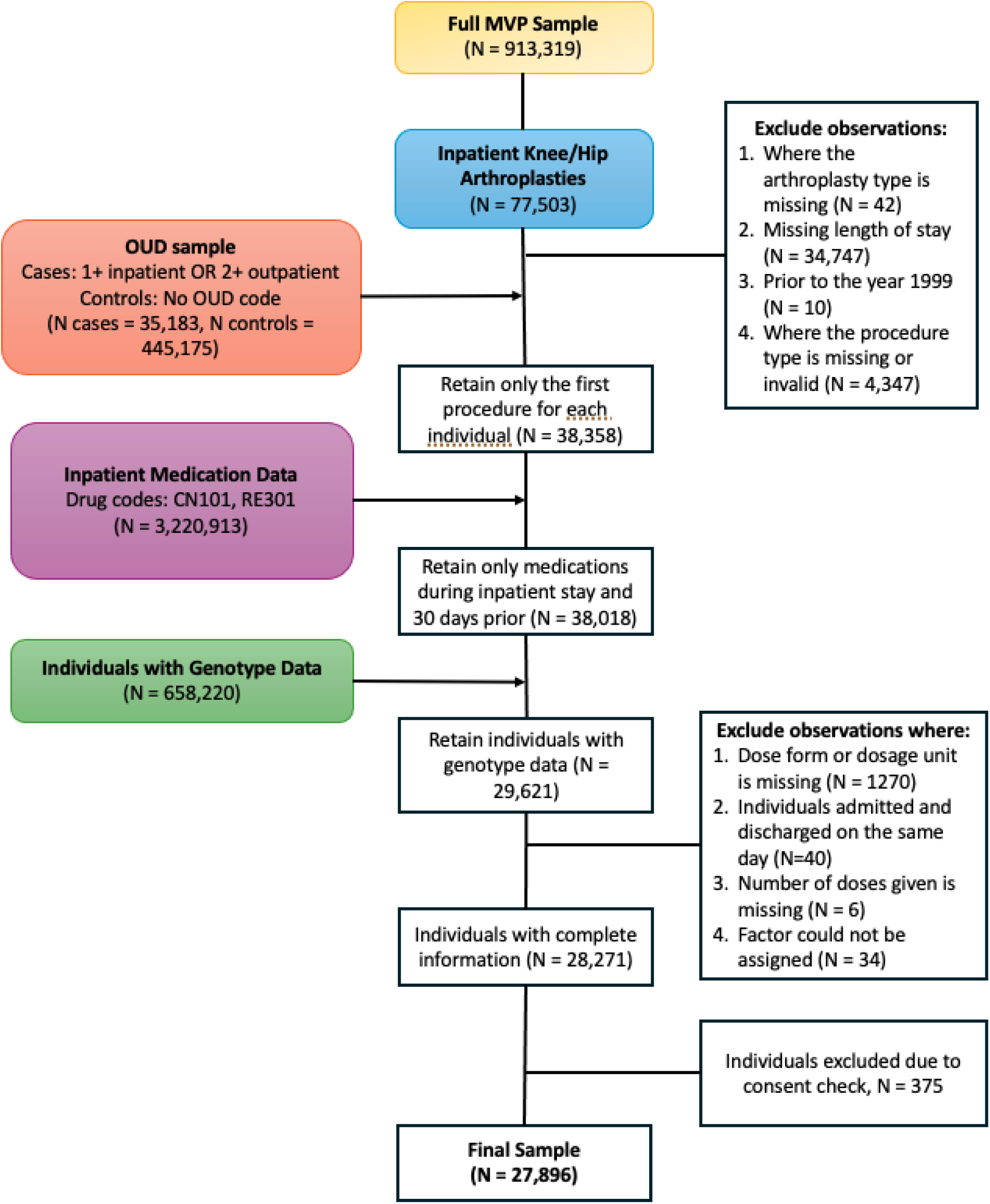
Schematic of the phenotype workflow

**Figure 2:**
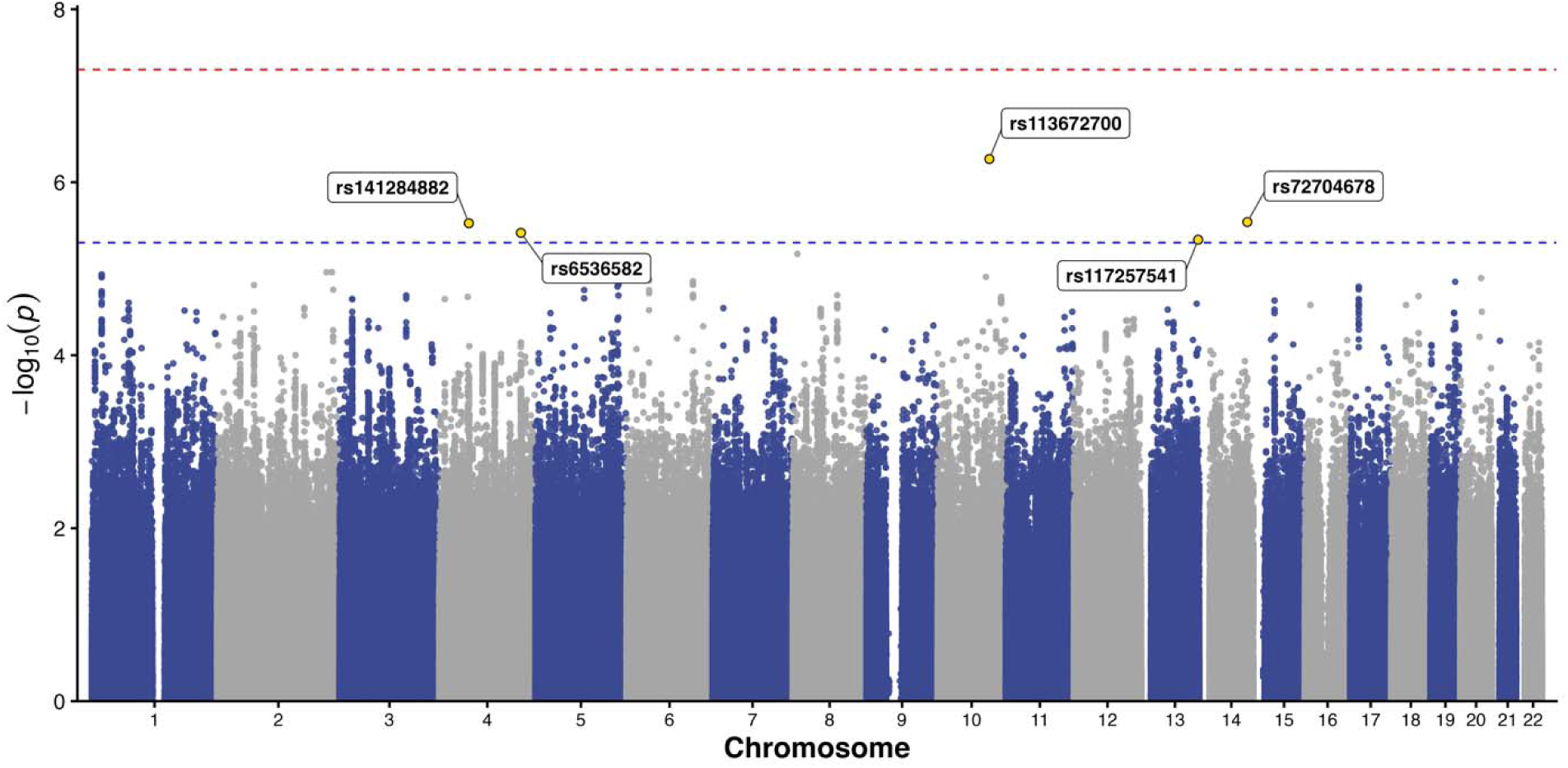
**Cross-ancestry genome-wide association results for opioid exposure following hip or knee arthroplasty.**

